# The epidemiological investigation of hypertension in northern Henan Province

**DOI:** 10.1101/2023.07.19.23292918

**Authors:** Xing Lu, Ziyang Lin, Junzheng Yang

## Abstract

**Background/purpose:** To understand the epidemiological status and risk factors of hypertension in northern Henan province, aim to provide a basis for the prevention, diagnosis, and treatment of hypertension in northern Henan Province.

**Methods:** To collect the clinical data in Xinxiang Central Hospital from 2019-2021, those data were classified and analyzed according to the basic information of investigation subjects including gender, ethnicity, age, smoking, marital status, occupation, education, and potential risk factors including hyperlipidemia, diabetes mellitus, ischemic stroke, coronary heart disease, angina pectoris, myocardial infarction, heart failure, peripheral artery disease, chronic renal insufficiency, gastrointestinal ulcers, chronic obstructive pulmonary disease, cerebral hemorrhage.

**Results:** A total of 770 clinical data of investigation subjects were collected, there are 621 males and 149 females, accounting for 80.65% and 19.35%, respectively; age range of investigation subjects was 20-97 years old; there were 248 males with hypertension, accounting for 32.21% in the total population, and there were 92 females with hypertension, accounting for 27.06% in the total population; there were 329 people with a history of smoking, and 346 people were never smoking, accounting for 49.10% and 51.64% in the total investigation population, respectively; in this investigation, the proportion of married people is the highest (638/670, 95.22%); the main occupational population was peasant farmer (353/670, 52.69%), followed by (164/670, 24.48%); the number of people with middle school/vocational school education was the highest(338/670, 50.45%), followed by primary school education (249/670, 37.16%).

the results of statistical analysis demonstrated that the incidence of hypertension is related to gender (P<0.01), hyperlipidemia (P<0.05), diabetes mellitus (P<0.001) and ischemic stroke (P<0.001), and had no relation to smoking, family history of coronary heart disease, family history of angina pectoris, history of heart failure, history of coronary intervention, history of peripheral arterial disease, chronic renal insufficiency, history of peptic ulcer disease, history of Chronic obstructive pulmonary disease and history of cerebral hemorrhage (P>0.05).

**Conclusion:** There is a certain regional specificity in the epidemiology of hypertension in the northern Henan Province, and the incidence of hypertension was related to gender, age, occupation, hyperlipidemia, diabetes mellitus and ischemic stroke, those evidences may provide clinical significance for the prevention, diagnosis, and treatment of hypertension in northern Henan province.

## Introduction

Hypertension is a clinical syndrome characterized by increased systemic circulation arterial blood pressure (systolic and/or diastolic) (systolic pressure≥103mmHg, diastolic pressure≥90mmHg) [1], and the first “Clinical Practice Guidelines for Hypertension in China” was released in 2022 which recommend to lower the diagnostic threshold of adult hypertension in China to systolic blood pressure≥130 mmHg and/or diastolic blood pressure≥80mmHg, hypertension usually accompanied by functional or organic damage to heart, brain, kidney and other organs [2]; especially, International Society of Hypertension in 2020 year reported that there was 54% stroke patients and 47% ischemic heart disease patient are caused by hypertension [3]. According to the investigation, there are about 8 million people die due to high blood pressure every year in the world [4, 5]. Hypertension is becoming the global health problem and causing a huge economic burden for individuals and countries due to its higher mortality rate and various complications, therefore, understanding the epidemiological status and related risk factors of hypertension is crucial for the prevention, diagnosis, and treatment of hypertension. For this purpose, we collected and analyzed the clinical data in Xinxiang Central Hospital from 2019-2021, hope to provide some evidences for hypertension treatment in north Henan province.

## Methods

A total of 770 clinical data were collected from in Xinxiang Central Hospital from 2019-2021, those data were classified and analyzed according to the basic information investigation subjects including gender, ethnicity, age, smoking, marital status, occupation, education, and risk factors including hyperlipidemia, diabetes mellitus, ischemic Stroke, coronary heart disease, angina pectoris, myocardial infarction, heart failure, peripheral artery disease, chronic renal insufficiency, gastrointestinal ulcers, chronic obstructive pulmonary disease, cerebral hemorrhage.

## Results

### 1 The correlation between general information of investigation subjects and hypertension in North Henan Province

Firstly, we analyzed whether the general information of investigation subjects including gender, ethnicity, age, smoking, marital status, occupation, and education level were correlated to the hypertension in north Henan Province. The results demonstrated that there were 340 hypertension patients including 248 male patients and 92 female patients in the total 770 clinical data; the age range of hypertension patients were 41-80 years old (286/340, 83.53%); there were 129 people were smoking (129/340, 37.94%), 326 people were married (326/340, 95.88%), the main occupation was peasant farmer (173/340, 50.88%), the main education level were primary school and middle school/vocational school (298/340, 87.65%) in 340 hypertension patients. The statistical results demonstrated that the gender, age and occupation were significantly correlated to the hypertension, and there were no correlation between hypertension and ethnicity, smoking, marital status and education level; especially, the people aged at 41-60 years old and the people aged at 61-80 years old had the high risk for hypertension (P=0.0171 and P=0.0304), and retirement person and professional technical personnel also had the high risk for hypertension (P=0.0152 and P=0.0355). Those data indicate that gender, age and occupation were the high risk factors for hypertension in north Henan Province, medical staff could take necessary prevention and diagnosis measures based on these risk factors.

**Table 1.** The correlation between general information of investigation subjects and hypertension in north Henan Province

| General information |  | Non-hypertensive (n=330) |  | Hypertensive (n=340) |  | Significance/<br>P value |
| --- | --- | --- | --- | --- | --- | --- |
|  |  | n | % | n | % |  |
| Gender | Male | 273 | 82.73% | 248 | 72.94% | 0.0029** |
|  | Female | 57 | 17.27% | 92 | 27.06% |  |
| Ethnicity | Han | 326 | 98.79% | 338 | 99.41% | 0.4446 |
|  | Hui | 4 | 1.21% | 2 | 0.59% | 0.4446 |
| Age | 20-40 | 17 | 5.15% | 6 | 1.76% | 0.0189* |
|  | 41-60 | 123 | 37.27% | 97 | 28.53% | 0.0171* |
|  | 61-80 | 153 | 46.36% | 187 | 55.00% | 0.0304* |
|  | ≥81 | 37 | 11.21% | 50 | 14.71% | 0.2062 |
| Smoking | Currently smoking | 139 | 42.12% | 129 | 37.94% | 0.2710 |
|  | Quit smoking ≥ 1 years | 18 | 5.45% | 23 | 6.76% | 0.5216 |
|  | Quit smoking ≥ 1 months | 8 | 2.42% | 7 | 2.06% | 0.7988 |
|  | Never smoking | 165 | 50.00% | 181 | 53.24% | 0.4395 |
| Marital status | Married (Remarried) | 312 | 94.55% | 326 | 95.88% | 0.4711 |
|  | Single | 2 | 0.61% | 1 | 0.29% | 0.6189 |
|  | Bereave | 13 | 3.94% | 11 | 3.24% | 0.6809 |
|  | Divorce | 3 | 0.91% | 2 | 0.59% | 0.682 |
| Occupation | Peasant farmer | 180 | 54.55% | 173 | 50.88% | 0.3539 |
|  | Worker | 53 | 16.06% | 49 | 14.41% | 0.5914 |
|  | Civil servants/<br>Government<br>officials | 2 | 0.61% | 6 | 1.76% | 0.2866 |
|  | Retirees | 67 | 20.30% | 97 | 28.53% | 0.0152* |
|  | Professional<br>technical personnel | 7 | 2.12% | 1 | 0.29% | 0.0355* |
|  | Individual business<br>owners | 4 | 1.21% | 3 | 0.88% | 0.7214 |
| Education<br>level | Illiteracy | 12 | 3.64% | 21 | 6.18% | 0.1538 |
|  | Primary school | 121 | 36.67% | 128 | 37.65% | 0.8108 |
|  | Middle<br>School/Vocational<br>School | 168 | 50.91% | 170 | 50.00% | 0.8172 |
|  | College/University | 28 | 8.48% | 20 | 5.88% | 0.2308 |
|  | Postgraduate | 1 | 0.30% | 1 | 0.29% | >0.9999 |

\*P<0.05, \*\*P<0.01.

### 2 The correlation between risk factors and hypertension in north Henan Province

And then we analyzed the correlation between hypertension and risk factors including hyperlipidemia, diabetes mellitus, ischemic stroke, coronary heart disease, angina pectoris, myocardial infarction, heart failure, peripheral artery disease, chronic renal insufficiency, gastrointestinal ulcers, chronic obstructive pulmonary disease, cerebral hemorrhage. The results demonstrated that hyperlipidemia, diabetes mellitus, ischemic stroke was highly correlated to the hypertension in north Henan Province, the P value was 0.0101, 0.0005 and 0.0008, respectively; there were no correlation between hypertension and the coronary heart disease, angina pectoris, myocardial infarction, heart failure, peripheral artery disease, chronic renal insufficiency, gastrointestinal ulcers, chronic obstructive pulmonary disease, cerebral hemorrhage (P>0.05).

**Table 2.** The correlation between risk factors and hypertension in north Henan Province

| Risk factors | Non-hypertensive<br>(n=330) |  | Hypertensive<br>(n=340) |  | Significance/<br>P value |
| --- | --- | --- | --- | --- | --- |
|  | n | % | n | % |  |
| Hyperlipidemia | 55 | 39.29% | 85 | 60.71% | 0.0101* |
| Diabetes mellitus | 47 | 14.24% | 85 | 25.00% | 0.0005*** |
| Ischemic Stroke | 34 | 10.30% | 67 | 19.71% | 0.0008*** |
| Coronary heart disease | 3 | 0.91% | 2 | 0.59% | 0.682 |
| Angina pectoris | 46 | 13.94% | 58 | 17.06% | 0.2868 |
| Myocardial infarction | 17 | 5.15% | 28 | 8.24% | 0.1239 |
| Heart Failure | 4 | 1.21% | 5 | 1.47% | >0.9999 |
| Coronary intervention | 9 | 2.73% | 17 | 5.00% | 0.1615 |
| Peripheral Artery Disease | 0 | 0.00% | 2 | 0.59% | 0.4994 |
| Chronic renal insufficiency | 2 | 0.61% | 2 | 0.59% | >0.9999 |
| Gastrointestinal ulcers | 9 | 2.73% | 7 | 2.06% | 0.6203 |
| Chronic Obstructive Pulmonary<br>Disease | 6 | 2.73% | 4 | 2.06% | 0.6203 |
| Cerebral hemorrhage | 2 | 0.61% | 5 | 1.47% | 0.451 |

\*P<0.05, \*\*\*P<0.0001.

## Discussion

Hypertension is a kind of chronic diseases characterized by elevated blood pressure, it can cause various subsequent diseases, seriously affecting the patient’s physical health and causing a huge burden [6, 7, 8, 9]. There are many factors could affect the occurrence and development of hypertension including genetic factors, environmental factors, age factors, habit factors, pharmacology factors, the impact of other diseases, which result in the complexity of hypertension prevention and treatment, therefore, early detection and prevention of hypertension are more important.

In this article, we investigated and analyzed the 770 clinical data were collected from in Xinxiang Central Hospital from 2019-2021, found that the general information including gender, age, and occupation were highly correlated to the hypertension in Henan Province; the number of male hypertension people were significantly bigger than female hypertension people (72.94% vs 26.06%); there is a high proportion of hypertension between the ages range of 40-80 years old, and retirement person and professional technical personnel had the higher risk for hypertension than that in peasant farmer and worker. Those data indicated that medical staff should pay special attention to male retirement person and professional technical personnel aged 40-80 years old and take necessary measures to decrease the probability of hypertension and reduce the harm caused by hypertension. Especially, many evidences demonstrated that smoking could enhance or result in hypertension directly or indirectly to threaten human health [10, 11, 12], but our data demonstrated that there is no correlation between smoking and hypertension (P>0.05), this may be because the number of participants in the survey was small, the specific reasons need the further research.

Subsequently, we analyzed the correlation between risk factors and hypertension found that hyperlipidemia, diabetes mellitus, ischemic stroke was highly correlated to the hypertension in North Henan Province, which is consistent with the results reported in many literature reports, this indicates that the occurrence of hypertension in northern Henan Province is consistent with the big data from “Clinical Practice Guidelines for Hypertension in China”, and relevant measures could be taken to reduce the incidence of hypertension.

## Conclusion

There is a certain regional specificity in the epidemiology of hypertension in the northern Henan Province, and the incidence of hypertension was related to gender, age, occupation, hyperlipidemia, diabetes mellitus and ischemic stroke, those evidences may help medical staff to prevention, diagnosis, and treatment of hypertension in northern Henan province.

## Data Availability

All data produced in the present study are available upon reasonable request to the authors

## Acknowledgements

None.

## References

[1] Daskalopoulou SS, Khan NA, Quinn RR, Ruzicka M, et al. The 2012 Canadian hypertension education program recommendations for the Management of Hypertension: blood pressure measurement, diagnosis, assessment of risk, and therapy. Can J Cardiol. 2012, 28:270–287.

[2] National Center for Cardiovascular Diseases; Chinese Medical Doctor Association; Hypertension Committee of the Chinese Medical Doctor Association; Chinese Society of Cardiology, Chinese Medical Association; Hypertension Committee of Cross-Straits Medicine Exchange Association, Chin J Cardiol, 2022, 50, 1050–1095 (In Chinese).

[3] Thomas Unger, Claudio Borghi, Fadi Charchar, Nadia A Khan, et al. A2020 International Society of Hypertension Global Hypertension Practice Guidelines. Hypertension. 2020, 75(6):1334–1357.

[4] American College of Sports Medicine. Guidelines for exercise testing and prescription. Philadelphia: Wolters Kluwer/Lippincott Williams & Wilkins Health; 2013.

[5] World Health Organization. Global Health Risks: Mortality and Burden of Disease Attributable to Selected Major Risks. Geneva; 2009.

[6] Di Palo KE, Barone NJ. Hypertension and Heart Failure: Prevention, Targets, and Treatment. Cardiol Clin., 2022, 40(2):237–244.

[7] Haseler E, Sinha MD. Hypertension in Children and Young Adults. Pediatr Clin North Am., 2022, 69(6):1165–1180.

[8] Lu Y, Chen R, Cai J, Huang Z, et al. The management of hypertension in women planning for pregnancy. Br Med Bull., 2018, 128(1):75–84.

[9] Buonacera A, Stancanelli B, Malatino L. Stroke and Hypertension: An Appraisal from Pathophysiology to Clinical Practice. Curr Vasc Pharmacol., 2019, 17(1):72–84.

[10] Sergey Dikalov, Hana Itani, Bradley Richmond, Aurelia Vergeade, et al. Tobacco smoking induces cardiovascular mitochondrial oxidative stress, promotes endothelial dysfunction, and enhances hypertension. Am J Physiol Heart Circ Physiol., 2019, 316(3): H639–H646.

[11] Fulvia Gloria-Bottini, M Banci, A Neri, A Magrini, et al. Smoking and hypertension: Effect of adenosine deaminase polymorphism. Clin Exp Hypertens., 2019, 41(6): 548–551.

[12] Zuxiang Wu, Yingxing Wu, Jingan Rao, Huan Hu, et al. Associations among vitamin D, tobacco smoke, and hypertension: A cross-sectional study of the NHANES 2001-2016. Hypertens Res., 2022, 45(12):1986–1996.

